# Rapid, Comprehensive Methylation-Based Classification of Hematologic Malignancies by Nanopore Sequencing

**DOI:** 10.64898/2026.07.02.26356825

**Authors:** Tristan Achterberg, Carlo Vermeulen, Huub van der Ent, Merel Jongmans, Karlijn Cammel, Esmée de Ruijter, Nicole Groenewegen, Cas Kranenburg, Marc van Tuil, Esmé Waanders, Mayur Parihar, Rubina Islam, Javeria Aijaz, Bianca Goemans, Friso Calkoen, Inge van der Sluis, Monique L. den Boer, Judith M. Boer, Valerie de Haas, Tim Triche, Thomas B. Alexander, Jeremy R. Wang, Nickhill Bhakta, Rob Pieters, Lennart Kester, Bastiaan Tops, Jeroen de Ridder

**Affiliations:** Princess Máxima Center for Paediatric Oncology; University Medical Center Utrecht, Center for Molecular Medicine; Oncode Institute; Princess Máxima Center for Paediatric Oncology; University Medical Center Utrecht, Center for Molecular Medicine; University Medical Center Utrecht, Department of Genetics; Tata Translational Cancer Research Center; Indus Health and Hospital Network; Princess Máxima Center for Paediatric Oncology; Center for Translational Immunology, University Medical Center Utrecht; Van Andel Institute; Department of Pediatrics, University of North Carolina; Department of Pathology and Laboratory Medicine, University of North Carolina; Department of Global Pediatric Medicine, Graduate School of Biomedical Sciences, St. Jude Children’s Research Hospital

## Abstract

Hematologic malignancies are diagnosed through a fragmented, sequential workup of morphology, immunophenotyping, cytogenetics, and molecular testing that can take days to weeks and is unavailable at many centers. DNA methylation profiling has transformed central nervous system tumor diagnosis, yet hematologic classifiers have remained confined to narrow acute leukemia panels. Here we present Lamprey, a deep-learning methylation classifier spanning 86 hematologic malignancy entities, trained on a reference cohort of 8,544 patients and deployed directly from nanopore sequencing. A depth-aware training framework allows confident classification from the first minutes of a run. Against blinded integrated reference diagnoses across retrospective, external, and prospective cohorts, Lamprey exceeded 98% accuracy among classified cases. Lamprey reaches a confident call within minutes, and cost as little as $82 per sample. Lamprey consolidates a sequential diagnostic workup into a single, rapid, same-day molecular readout.

## Introduction

Hematologic malignancies comprise a biologically diverse group of clonal disorders whose accurate and timely classification determines treatment. Diagnosis currently depends on the sequential integration of morphology, flow cytometry, immunohistochemistry, cytogenetics, fluorescence in situ hybridization (FISH), and targeted or genome-wide sequencing.^1–4^ Because the disease category is usually unknown at presentation, these assays are applied iteratively, each result narrowing the differential and dictating the next, so that a definitive diagnosis is often reached only after days to weeks. Diagnostic delay or misclassification carries direct clinical consequences: subtypes defined by cryptic or non-canonical fusions (such as *DUX4*-rearranged, *BCR*::*ABL1*-like, and *NUP98*-rearranged leukemias) can be missed without whole-genome or transcriptome sequencing,^5^ lineage-ambiguous leukemias resist stable immunophenotypic classification, and overlapping lymphoid entities generate substantial interobserver variability.^6,7^ This burden falls hardest on centers without access to the full diagnostic armamentarium, where many patients never receive a complete molecular diagnosis.

DNA methylation profiles encode both the cell of origin and the alterations acquired during malignant transformation.^8^ Methylation-based machine learning classifiers have transformed the diagnostic workup of central nervous system tumors by resolving morphologically ambiguous cases and identifying biologically meaningful subgroups^9–11^. This approach has been extended to intraoperative care, with rapid nanopore sequencing enabling real-time methylation-based classification during neurosurgery and providing actionable information within the timeframe of a surgical procedure^12–14^. In hematologic malignancies, however, this approach has been confined to classifiers covering a limited set of acute leukemia subtypes and validated in small, single institution, cohorts, leaving the broader and most diagnostically challenging entities unaddressed.^15–17^

We developed Lamprey, a DNA methylation–based classifier spanning 86 entities across the full spectrum of hematologic malignancies (acute and chronic leukemias, myelodysplastic syndromes, lymphomas, plasma-cell neoplasms, and histiocytic disorders), built from the largest methylation reference cohort assembled for these diseases to date. Model development required confronting two obstacles that have constrained methylation-based classification more broadly: label noise in retrospective reference cohorts, and the systematic distortion that arises when array-trained models are applied to nanopore data. Lamprey classifies samples directly from nanopore sequencing and can return a confident result within minutes of a run, allowing for rapid turnaround times. This speed changes how a diagnostic workup can be sequenced. Rather than committing every sample to the full cascade of assays up front, a single nanopore run can triage in real time: the majority of cases reach a confident classification within the first hour, while cases that remain below the confidence threshold are flagged for resolution by the copy-number, structural-variant, and sequence-variant data accumulating on the same run, rather than misassigned. We validated Lamprey in retrospective and prospective patient cohorts, in independent collaborator cohorts and additionally assessed time-to-result and per-sample cost as implementation outcomes to determine feasibility of realistic deployment scenarios. Because the same data can also be used to identify copy-number profiles, structural variants, and somatic variants (validated in a study of the nanopore variant caller, NASVar^18^), methylation classification and DNA alteration data are consolidated into a single assay, replacing what currently requires multiple sequential tests.

## Methods

### Study design and cohorts

We trained Lamprey on a DNA methylation reference cohort of 8,544 patients (9,000 samples) spanning 86 diagnostic entities, assembled from 84 published microarray datasets and in-house data (Supplementary Data Table 1). Diagnostic accuracy was evaluated in three independent nanopore cohorts that were not used in any aspect of model development: a retrospective cohort of 378 patients (380 samples; 291 pediatric and 87 adult), assembled from in-house nanopore data (Princess Máxima Center and UMC Utrecht) and published datasets and enriched for rare and diagnostically challenging entities; three independent pediatric acute leukemia cohorts (244 patients) from collaborating institutions (UNC/St. Jude; the Tata Translational Cancer Research Center [TTCRC], Kolkata; and The Indus Hospital, Karachi, Pakistan); and a prospective cohort of 18 consecutively accrued patients at the Princess Máxima Center, sequenced on the day of or the day after sample receipt.

### Definition of methylation classes

Class definitions followed the 2022 World Health Organization (WHO) classification^19,20^ except where methylation data supported a biologically meaningful refinement. The methylation classes *NPM1* with *IDH* mutation, TET2 mutation or DNMT3A mutation, *KMT2A*-rearranged acute myeloid leukemia (AML) with *MECOM*, *HMX3*, *FOXC1* or *HOXA*/*B* activation, and AMKL-mixed were adopted as defined in Steinicke et al.^17^ Samples with FET-ETS family fusions such as *FUS*::*ERG*, *FUS*::*FLI1*, *FUS*::*FEV* or *HNRNPH1*::*ERG* were combined. *ETV6*::*RUNX1* and *ETV6*::*RUNX1*-like were merged into one class. High-hyperdiploid, low-hypodiploid and near-haploid B-cell precursor acute lymphoblastic leukemia (BCP-ALL) were merged into a BCP-ALL with aneuploidy class.

Juvenile myelomonocytic leukemia (JMML)-low, -medium and -high methylation classes were reproduced as in Schönung et al.^21^ Myelodysplastic syndrome (MDS) was stratified into hMDS (high International Prognostic Scoring System [IPSS] risk, *TP53* mutation, aneuploidy) and lMDS, with *IDH*-mutant cases forming a separate class. The T-cell acute lymphoblastic leukemia (T-ALL) classes *TAL1*-like, early T-cell precursor (ETP)-enriched and T-ALL other were derived from Touzart et al.^22^, with *TLX1*- and *TLX3*-enriched groups merged into T-ALL other. Langerhans cell histiocytosis and Erdheim-Chester disease were merged. Diffuse large B-cell lymphoma (DLBCL) and primary mediastinal large B-cell lymphoma (PMBCL) were merged owing to lack of PMBCL training data. Mantle cell lymphoma was separated by *SOX11* expression, chronic lymphocytic leukemia (CLL) by *IGHV* mutation status, and Burkitt lymphoma by Epstein–Barr virus (EBV) status, with high-grade B-cell lymphoma with 11q alterations merged with EBV-negative Burkitt lymphoma. Extranodal T-cell lymphoma NOS was divided into *SETD2*-altered and non-*SETD2* subgroups.

### Reference standard

The reference diagnosis was established independently of Lamprey’s methylation classification by routine integrated hematopathology, including morphology, flow cytometry, cytogenetics, FISH, immunohistochemistry, and clinically indicated molecular testing; in a subset of cases, it incorporated sequence-variant and structural-variant calls derived from the same nanopore run (NASVar). Treating clinicians and diagnostic laboratories were blinded to Lamprey’s methylation-based predictions when assigning the reference diagnosis.

Because Lamprey classifies on DNA methylation, an orthogonal readout to sequence-variant calling, its predictions were derived from data features independent of those informing the reference diagnosis.

### Nanopore sequencing

Mononuclear cells were isolated from peripheral blood or bone marrow aspirates by Ficoll density-gradient centrifugation; for lymphoma samples, genomic DNA was extracted from diagnostic tissue biopsies. Genomic DNA was isolated with the AllPrep DNA/RNA Kit (Qiagen) on a QIAcube platform and quantified by Qubit fluorometry. For each library, 1 µg of DNA was fragmented with Covaris g-Tubes, repaired and end-prepared with the NEBNext FFPE DNA Repair Mix and Ultra II End Prep Kit (New England Biolabs), and prepared with the Oxford Nanopore Technologies Ligation Sequencing Kit v14; the Native Barcoding Kit v14 was used for multiplexed samples. Libraries were sequenced on R10.4.1 flow cells (FLO-PRO114M) on a PromethION P2i or P24 instrument, with real-time high-accuracy basecalling and simultaneous detection of 5-methylcytosine and 5-hydroxymethylcytosine in CpG context. Adaptive sampling against a custom reference and target-region set was enabled to enrich loci relevant to leukemia classification.^18^

### Classification and confidence threshold

Methylation profiles were classified by an ensemble of ten deep neural networks trained to operate across the full range of sequencing depths encountered in clinical practice. Model confidence was calibrated such that the prespecified threshold of 0.90 corresponds to a high probability of a correct classification; samples below this threshold were not assigned a classification.

### Simulation of sparse nanopore profiles

Because Lamprey is trained on methylation-array profiles but deployed on nanopore sequencing, each array profile was used to generate simulated nanopore observations across training iterations. For each iteration, the number of covered CpG sites was sampled between 5,000 and 353,232 using inverse square-root–weighted sampling. For each covered probe, the per-site read depth (Nvalid) was drawn from a lognormal distribution whose mean and standard deviation (μ and σ in log-space) were modelled as smooth spline functions of sample depth, estimated from 733 empirical nanopore samples (R² = 0.97); at simulation time, μ and σ were evaluated at the sampled depth, perturbed by draws from depth-stratified residual distributions, and used to generate per-site Nvalid values. To reproduce the systematic bias of the nanopore methylation caller, the probability of a positive call was modelled as a logistic function of the array beta value, P(call = 1 | β) = σ(a + b·logit(β)); parameters fitted under a binomial likelihood (a = −0.155, b = 1.517) were stable across read-depth strata, indicating that caller bias is independent of depth. A nanopore methylation call was then simulated by drawing a binomial count of modified reads from Binomial (Nvalid, p_eff), where p_eff is the caller-bias-corrected effective methylation probability, and computing the resulting fractional methylation. Calls were binarized: fractional methylation above 0.5 was encoded as 1, below 0.5 as −1, exactly 0.5 as 0; uncovered sites were encoded as 0.

### Classifier architecture and training

Lamprey is a residual multilayer perceptron that takes 353,232 CpG features through a projection to 512 dimensions, two residual blocks at 512 dimensions, a projection to 256 dimensions with one residual block, and a projection to 128 dimensions before the classification layer; each residual block applies a linear layer, layer normalization, dropout (0.5), and a SiLU activation with a skip connection. Training used the AdamW optimizer (learning rate 0.001 with 5,000-step warmup and cosine annealing to 0.0001; weight decay 0.01) for 50 epochs of 100,000 class-balanced instances each. Ten networks were trained under grouped, stratified five-fold cross-validation, with groups defined by patient and diagnostic class and one fixed held-out test fold, and their per-class scores were averaged to form the ensemble prediction.

### Confidence calibration

To address systematic miscalibration of model confidence as a function of sequencing depth, we applied post-hoc temperature scaling calibrated on the held-out microarray test set. For each sample, the depth-aware simulation framework was used to generate nanopore-like inputs across the full range of CpG coverage, and ensemble-averaged logits were computed from all ten models. Logits were aggregated into adaptive CpG coverage bins with finer resolution at low coverage (1,000-step increments below 20,000 CpGs, 2,000-step increments from 20,000 to 40,000, 5,000-step increments from 40,000 to 80,000, 10,000-step increments from 80,000 to 110,000, and 20,000-step increments above 110,000 CpGs) to capture the more rapid changes in calibration behavior at low sequencing depths. A separate scalar temperature parameter T was optimized independently per bin by minimizing cross-entropy loss on the aggregated logits using the L-BFGS optimizer with the strong Wolfe line search condition. Temperature scaling operates by dividing the raw logits by T, where T > 1 softens the predicted probability distribution (correcting overconfidence) and T < 1 sharpens it (correcting under confidence). Calibration performance was assessed using expected calibration error (ECE), computed across 10 equal-width confidence bins as the weighted mean absolute difference between mean confidence and accuracy within each bin. At inference time, the appropriate temperature is selected based on the observed CpG coverage of the input sample.

### Dataset curation and label propagation

Our originally defined dataset contained 6518 labelled samples out of 9000 total samples. As diagnoses may not have been captured by existing experimental techniques or were not considered, we employed a filtering strategy. We trained a residual neural network on 200,000 CpG sites to obtain highly accurate scores on our original dataset. 5 residual models were trained using a 5-fold cross-validation scheme, with 20% of the data being excluded as the test set for each model. Dimensionality reduction was performed using Principal Component Analysis (PCA) on the top 50,000 most variable CpG sites, retaining the first 100 principal components. For each sample, the twenty nearest neighbors were identified in PCA space and label concordance among these neighbors was calculated. Labels were removed if samples had one or less concordant label among its neighbors, and the label received a prediction of 0.05 or less from the model within the 5-fold cross-validation. The 5-fold cross-validation was rerun on the cleaned dataset and then used all five models to predict on 50 simulated nanopore runs per unlabeled sample. Samples achieving a mean confidence ≥0.95 across all folds and simulations were assigned the highest-predicted class. All nanopore validation cohorts were analyzed as independent test sets and were not subjected to Lamprey-informed exclusion, cleaning, or relabeling.

### Benchmarking against prior classifiers

To compare Lamprey against previously published methylation-based classifiers, we benchmarked against ALMA v2 and MARLIN using acute leukemia samples from the respective published validation cohorts supplemented with in-house nanopore data. For each classifier, we applied the confidence thresholds described in the original publications (≥0.5 for ALMA, ≥0.8 for MARLIN). For samples from the Princess Maxima Center cohort, modkit pileup was rerun using the parameters and probe feature sets specified on the respective tool GitHub repositories.

Because ALMA, MARLIN, and Lamprey operate on different class taxonomies, we established equivalence rules to ensure a fair comparison. For MARLIN, a prediction was scored as correct when the predicted methylation group encompassed the true entity: Ph/Ph-like was accepted for both *BCR*::*ABL1*-positive and *BCR*::*ABL1*-like BCP-ALL; *TCF3*::*PBX1*/*MEF2D* was accepted for either *TCF3*::*PBX1* or *MEF2D*-rearranged BCP-ALL; and HOX group 4 was accepted for *NPM1*-mutated, *UBTF*-ITD, *NUP98*-rearranged, *DEK*::*NUP214*-rearranged, or *KMT2A*-rearranged AML. These merged predictions were scored as correct for MARLIN, meaning that Lamprey’s accuracy is measured at a finer diagnostic resolution than MARLIN’s throughout the comparison. ALMA distinguishes high hyperdiploidy from hypodiploidy and near-haploidy as separate classes; predictions within this group were therefore scored strictly. MARLIN does not separate these entities, and predictions within the aneuploid group were scored as correct regardless of subtype.

### Statistical analysis

The primary measures were diagnostic accuracy among classified samples (concordance with the reference diagnosis) and diagnostic yield (the proportion of samples receiving a confident classification). Binomial 95% confidence intervals (CIs) for accuracy and diagnostic yield were calculated by the Wilson method.

### Time-to-diagnosis analysis

For single-plex runs (n=174), in which one sample received the full output of a flow cell, per-read sequencing timestamps were extracted from the sequencing output files and reads were cumulatively included by elapsed sequencing time. At successive time points, modkit pileup was recomputed over all reads generated up to that point and the profile was classified by the ten-model ensemble with coverage-dependent temperature scaling. The time to confident classification was the earliest time point at which the calibrated top-class confidence reached 0.90. Per-sample and median trajectories were computed, and the cumulative proportion reaching threshold tabulated at 10, 30, 120, and 400 minutes and at run completion. A six-plex configuration was approximated by rescaling the single-plex time axis by a factor of six, reflecting the division of one flow cell’s output across six barcoded samples (Extended Data Fig. 6).

### Cost analysis

Per-sample reagent and consumable costs were estimated by partial microcosting, following the framework developed for the companion NASVar analysis, spanning DNA extraction, library preparation, quality control, and sequencing, grouped for presentation into flow cell; library preparation; and reagents and consumables. Activity-based costs (instrument capital, personnel time, and computational infrastructure) were excluded as sunk costs, on the basis that the nanopore assay displaces rather than adds to existing conventional pathology workflows. Costs were modelled under three throughput scenarios: 6-plex, 12-plex, and 24-plex native barcoding, with flow-cell and wash costs amortized across multiplexed samples and per-sample library and consumable costs held constant. For each line item, low and high estimates were taken as the minimum and maximum procurement prices observed across three participating centers spanning high- and low/middle-income settings (The Indus Hospital; TTCRC Kolkata; UNC).

### Data availability

Pediatric patient data from the Princess Máxima Center are currently being prepared for submission to GEO and the European Genome-Phenome Archive (EGA) and will be made available under accession numbers xxxx.

### Code availability

Code for the Lamprey prediction tool is available as a Python package and can be found at: https://github.com/princessmaximacenter/Lamprey

### Patient identity

Patient identifiers for samples obtained from the Princess Máxima Center biobank were already pseudonymized prior to access. The link between the provided identifiers and patient identities is not accessible outside the originating biobank. For publicly available datasets, the original identifiers provided in the repository metadata were retained.

## Results

### A reference cohort spanning the hematologic malignancy spectrum

We assembled the largest DNA methylation reference cohort for hematologic malignancies to date, comprising 8,544 patients across 86 entities spanning acute and chronic leukemias, myelodysplastic syndromes, lymphomas, plasma-cell neoplasms, and histiocytic disorders (Fig. 1A–C). Class boundaries were refined from the 2022 WHO classification baseline where methylation data supported a biologically meaningful distinction (Extended Data Fig. 1, 2C,D). We prioritized clinical actionability over unsupervised cluster purity. Where entities are genetically and clinically distinct yet converge on methylation, we kept them separate rather than collapsing them by epigenetic similarity, as prior classifiers have done^17^; for example, NPM1-mutated and KMT2A-rearranged AML, which share a HOXA-activated methylation program but differ in risk profile, are retained as separate classes (Extended Data Fig. 3D). A small number of entities could not be reliably separated by methylation-based classification alone, either due to overlapping methylation profiles or lack of training data of one subtype, and were grouped (Extended Data Fig. 1, 2C,D). In some of these cases the distinguishing lesion is recoverable from the same sequencing run by copy-number and structural-variant analysis. Mixed-phenotype acute leukemias were merged with their molecularly corresponding subtypes because their methylation profiles are indistinguishable from those of the genetically defined entity sharing the same driver lesion (Extended Data Fig. 3A).^23^ The resulting taxonomy is substantially broader than prior methylation-based classifiers, which have focused on acute leukemias or single disease categories, and enables classification across diagnostic categories without prior lineage stratification.

**Figure 1.**
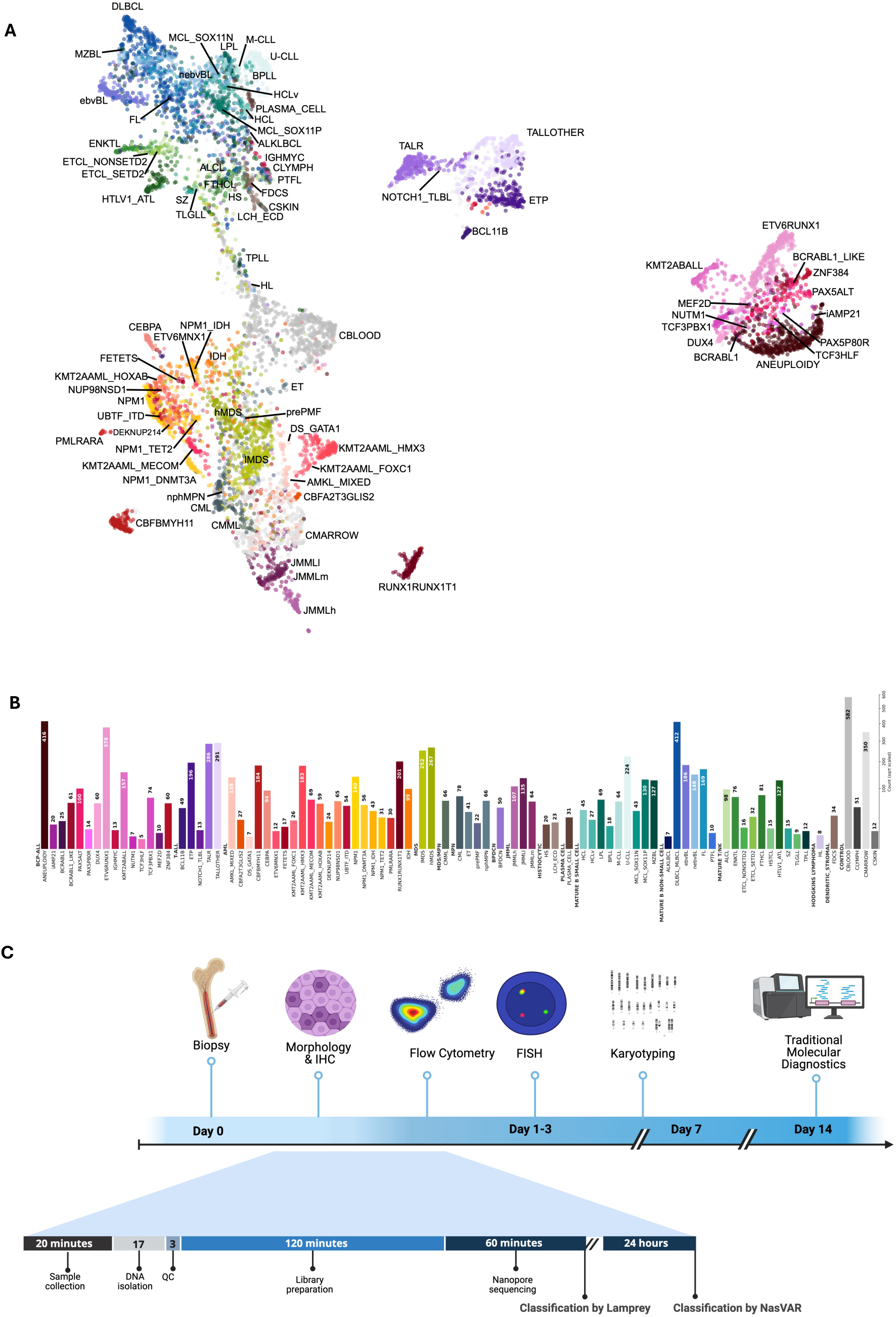
Methylation-based classification across hematologic malignancies and the nanopore diagnostic workflow. **A.** UMAP of DNA methylation profiles from 8,544 hematological malignancy patients, colored by methylation class (n=86) and labeled with the class abbreviation. Labels were assigned through a two-stage cleaning pipeline combining neural-network confidence filtering and PCA-KNN neighborhood concordance, followed by label propagation for unlabeled public samples. **B.** Sample counts per methylation class in the final Lamprey training dataset, grouped by disease family (square-root-scaled x-axis). **C.** Top: standard-of-care diagnostic workflow from biopsy to final diagnosis, including sequential histopathological, flow-cytometry, and molecular testing steps. Bottom: diagnostic timeline incorporating nanopore sequencing into routine care, demonstrating rapid turnaround time.

Visualizing the cohort by uniform manifold approximation and projection (UMAP) showed clear lineage-based separation of myeloid leukemias, B-cell precursor ALL, T-ALL, and mature lymphoid neoplasms (Fig. 1A), but it also exposed a tension between methylation-based clustering and established genetic classification. Genetically distinct but biologically related entities, such as *HOXA*-activated myeloid leukemias, co-cluster on UMAP despite markedly different prognoses and therapeutic implications. Crucially, overlap in a two-dimensional embedding does not imply that these entities are inseparable in the full feature space: a classifier operating on hundreds of thousands of CpG sites can exploit subtle but consistent differences that a low-dimensional projection necessarily obscures. We therefore used UMAP co-clustering only to resolve leukemias of ambiguous origin, and otherwise preserved genetically and clinically defined boundaries rather than collapsing entities by epigenetic similarity.

### Curating a high-quality reference cohort under label noise

Label noise in retrospective reference annotations is a central but rarely addressed obstacle to accurate methylation-based classification. Diagnoses may reflect outdated classification schemes, ambiguous immunophenotyping, or incomplete molecular workup at the time of collection. We therefore applied a two-stage pipeline combining confident-learning–based label cleaning with semi-supervised label propagation, the latter resolving 70 of 91 unlabeled mixed-phenotype acute leukemia samples into their molecular subtypes (Extended Data Fig. 2A,B, 3B).^24–26^ On held-out nanopore cohorts, curation raised the proportion of confident classifications from 51.8% to 71.4% without loss of accuracy (Extended Data Table 1). Because reference annotations inevitably age as molecular classification is revised, this form of label noise is intrinsic to any methylation classifier assembled from historical cohorts rather than peculiar to this dataset; the curation procedure is agnostic to the disease classes themselves and remains applicable as reference sets continue to grow. Propagation also recovered known biology: T-cell acute lymphoblastic leukemia (T-ALL) samples annotated only by CpG island methylator phenotype (CIMP) status were assigned molecular subtypes coherently (Extended Data Table 2). CIMP-negative cases mapped predominantly to the TAL-like subtype, an association linked to minimal residual disease–defined poor-risk disease.^27,28^

### Lamprey achieves high performance on a simulated nanopore test set

In five-fold cross-validation across simulated nanopore coverage, Lamprey achieved a balanced accuracy of 0.91 and an overall accuracy of 0.93, rising to 0.96 and 0.98, respectively, among samples meeting the 0.90 confidence threshold (Fig. 2A and 2B). Confidence was well calibrated after coverage-dependent temperature scaling, which reduced the expected calibration error from 0.0157 to 0.003 (Fig. 2C). Rare subtypes with few training samples were variably resolved (e.g., *NUTM1*-rearranged BCP-ALL, F1 = 0.62; Down syndrome–associated *GATA1*-mutant AML, F1 = 0.90). Entities deprecated in the 2022 WHO classification, such as B-cell prolymphocytic leukemia and hairy-cell leukemia variant, nonetheless retained methylation signatures distinct enough for classification.

**Figure 2.**
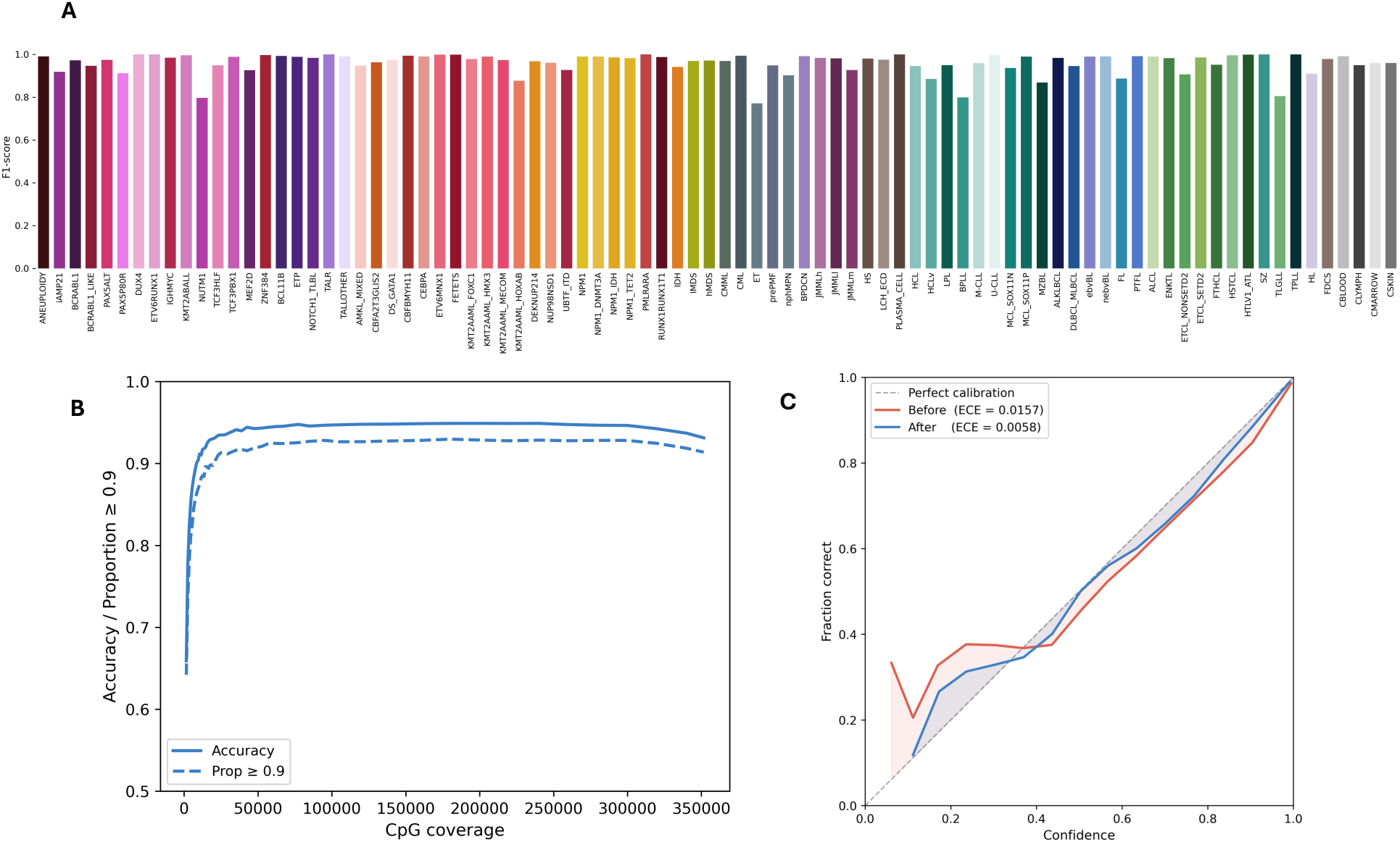
Cross-validated classification performance and calibration. **A.** Per-class F1 scores from 5-fold cross-validation on the final Lamprey training dataset (86 classes), restricted to samples with 50,000-100,000 covered CpG sites and a classification score of ≥0.9. **B.** Classification accuracy and proportion of samples exceeding a classification score of ≥0.9 as a function of CpG site coverage, reflecting expected performance across a range of nanopore sequencing depths. **C.** Calibration plot comparing predicted classification scores to observed class frequencies before and after bin-based temperature scaling. A perfectly calibrated model follows the diagonal (dashed line).

### A depth-aware simulation and residual architecture jointly raise accuracy and confidence

Sequencing depth in clinical nanopore runs is neither fixed nor predictable, varying with run duration, sample quality, and clinical urgency, yet prior methylation classifiers were each trained at a single coverage regime.^12,16,17,29^ A classifier trained at a fixed depth learns to rely on statistical regularities specific to that coverage level, which can mean poor generalization when deployed at a sequencing depth that is disconnected from its training distribution. We therefore developed a simulation framework that, for the first time, generates nanopore observations from distributions fitted to real runs. We therefore drew both the number of covered CpG sites and the per-CpG read depth from empirical models rather than by uniform downsampling (Extended Data Fig. 4), paired with an architecture able to exploit that variability.

Replacing the original feedforward architecture with the residual network and training across the full depth-aware simulation, rather than at sparse fixed coverage or with uncorrected Bernoulli sampling, jointly raised held-out accuracy to 87.5% and the confident-call rate to 71.4%. Removing either component lowered both accuracy and model confidence (Extended Data Table 3). The depth-aware simulation in turn depended on an explicit model of the nanopore methylation caller: matched array and nanopore profiles revealed a systematic, S-shaped bias in per-site calls that was independent of read depth and was corrected by a fitted logistic transfer function before training (Extended Data Fig. 4A). Modelling this bias, rather than treating array beta values as direct methylation call probabilities, was necessary to close the array-to-nanopore domain gap. Because the distortion originates in the basecaller itself rather than in any property of Lamprey, it is inherited by any classifier that ports array-trained methylation models onto nanopore data; characterizing it directly is what makes training across the full range of clinically encountered coverage possible.

### Evaluation on a retrospective nanopore cohort

In the retrospective cohort (380 samples, 53 entities), Lamprey classified 244 of 380 samples (64.2%; 95% CI, 59.3 to 68.9) at the 0.90 threshold, with an accuracy of 98.4% among classified cases (240 of 244; 95% CI, 95.9 to 99.4) (Fig. 3A). Unclassified samples were concentrated in low-tumor-purity entities: Langerhans-cell histiocytosis and Hodgkin’s lymphoma reached the threshold in only 7 of 61 cases, and excluding these entities raised the classification rate to 74.3%. Both are characterized by low tumor-cell content; although they classify confidently on methylation arrays (85.3% and 88.1%, respectively), on nanopore data the combination of low purity and residual platform domain shift may push them below the confidence threshold. In serial-dilution experiments, classification confidence declined below approximately 50% tumor content (Extended Data Fig. 5). Accuracy was similar in pediatric and adult samples (98.4% vs. 98.3%).

**Figure 3.**
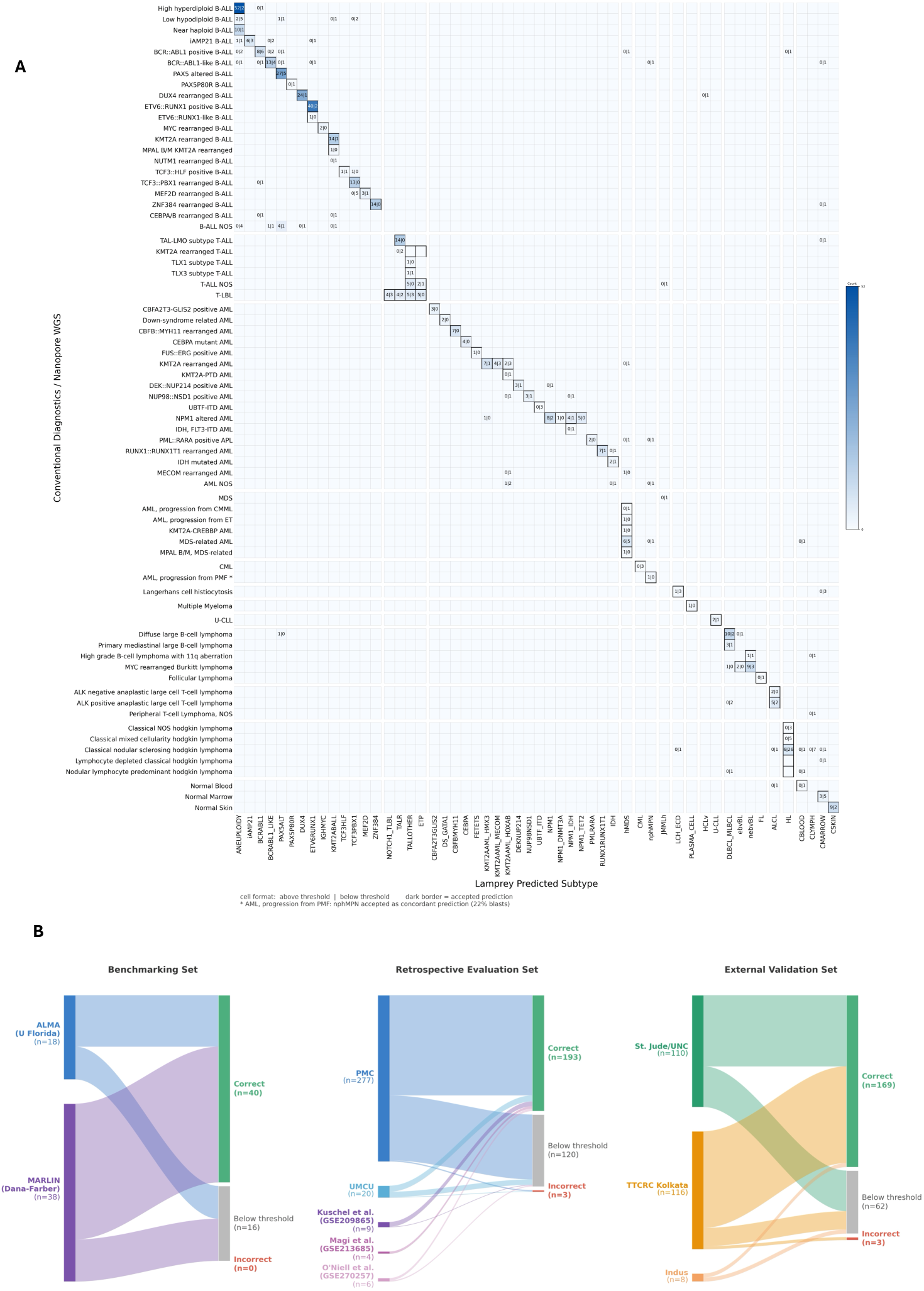
Classification accuracy across nanopore cohorts. **A.** Confusion matrix of Lamprey predictions on all nanopore cohorts. Rows represent sample diagnoses, as determined by conventional diagnostic workflows (FISH, RNA-seq, WGS, flow cytometry, etc.) or NASVar; columns represent predicted labels. Cell values show the number of samples classified above (≥0.9) and below (<0.9) the confidence threshold, separated by a vertical bar. Predictions considered correct are highlighted by a darkened cell border. Asterisk: AML, progression from PMF — nphMPN accepted as concordant prediction (22% blasts). **B.** Sankey diagram showing Lamprey classification results on the benchmarking, retrospective evaluation, and external validation cohorts. Samples are stratified by whether they reach the 0.9 confidence threshold; those above threshold are labeled correct or incorrect. Samples with an inconclusive reference diagnosis were omitted.

### Methylation-based classification captures disease biology beyond categorical boundaries

Lamprey’s classifications tracked underlying disease biology rather than immunophenotype. Two mixed-phenotype acute leukemias, whose surface immunophenotype is ambiguous, were each classified with high confidence into the molecular subtype matching their driver alteration (a *KMT2A*-rearranged case as *KMT2A*-rearranged BCP-ALL, and a myelodysplastic syndrome (MDS)-related case as high-risk MDS). A primary myelofibrosis sample with 22% blasts retained its identity as a non-Philadelphia myeloproliferative neoplasm despite approaching the threshold for transformation, whereas an essential thrombocythemia in blast phase (21.8% blasts) was classified as high-risk MDS/secondary AML, consistent with epigenetic reprogramming having occurred in the latter but not the former. Although demethylating agents broadly reduce DNA methylation, the features that distinguish subtypes appeared to persist under such treatment. A sample obtained at relapse during azacitidine treatment from a patient with KMT2A-rearranged AML was classified correctly with high confidence (ALMA patient 11), and high-risk or secondary AML samples profiled before and after demethylating therapy co-localized on a uniform manifold approximation and projection (UMAP) embedding regardless of treatment (Extended Data Fig. 3C).

### Validation in independent external cohorts

In three independent pediatric acute leukemia cohorts from collaborating institutions (244 patients; UNC/St. Jude, TTCRC Kolkata, and The Indus Hospital, Pakistan), Lamprey classified 176 of 244 samples using the diagnostic threshold of 0.9 (72.1%; 95% CI, 66.2 to 77.4); among classified cases with a definitive reference subtype, accuracy was 98.3% (169 of 172; 95% CI, 95.0 to 99.4) (Fig. 3B).

### Lamprey outperforms existing methylation-based acute leukemia classifiers

We compared Lamprey with the existing methylation-based classifiers ALMA v2^16^ and MARLIN^17^, restricted to acute leukemia samples. Accuracy was assessed only among samples exceeding each classifier’s predefined reporting threshold. Lamprey achieved higher accuracy (98.8% vs. 96.1% for ALMA v2 and 86% for MARLIN) while classifying a higher proportion of samples than ALMA v2 (75.5% vs. 59.7%); MARLIN reached its reporting threshold in 86.1% of samples. It assigned diagnoses across entities absent from ALMA (including *DUX4*- and *ZNF384*-rearranged BCP-ALL and *UBTF*-ITD AML) and separated entities that MARLIN groups together, including the menin-inhibitor–susceptible AML categories (*NPM1*-mutated, *DEK*::*NUP214*-, *UBTF*-ITD, *KMT2A*-, and *NUP98*-rearranged) and *BCR*::*ABL1* versus *BCR*::*ABL1*-like BCP-ALL (Fig. 4A, B).

**Figure 4.**
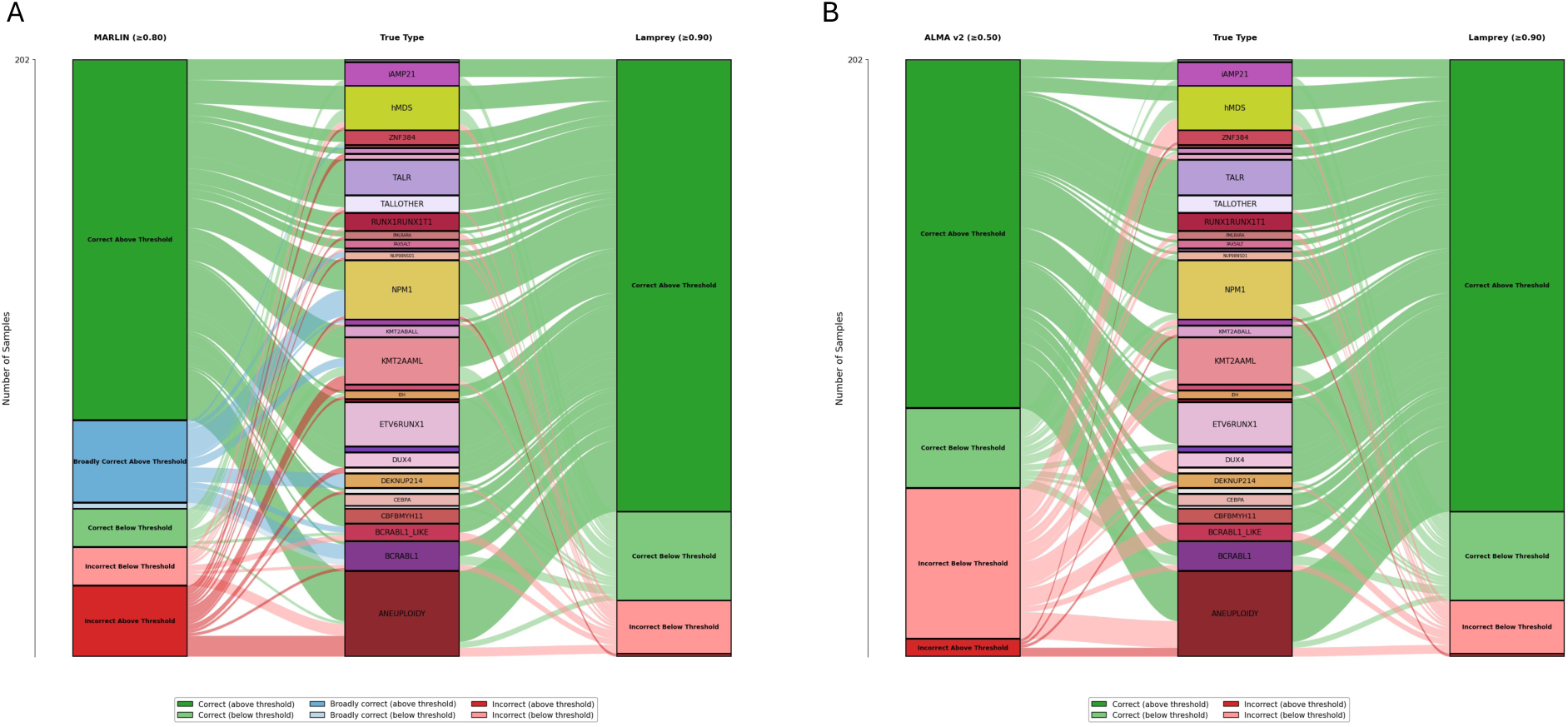
Comparison of Lamprey with prior methylation-based classifiers on acute leukemia. **A.** Alluvial plot comparing MARLIN predictions at its recommended 0.8 confidence threshold (left) with Lamprey predictions at the 0.9 threshold (right) on the retrospective nanopore cohort (Princess Máxima Center, University of Florida, and Dana-Farber Cancer Institute), with reference diagnoses in the center column. Flows and terminal boxes are colored by classification outcome: correct, broadly correct (the prediction encompasses the true entity within a broader diagnostic group), or incorrect, each stratified by whether the prediction reached the confidence threshold. **B.** Alluvial plot comparing ALMA v2 predictions at its recommended 0.5 confidence threshold (left) with Lamprey predictions (right) on the same cohort; because ALMA’s taxonomy does not include merged diagnostic groups, only correct and incorrect outcomes are distinguished.

This difference likely reflects Lamprey’s depth-aware simulation framework: ALMA v2 was trained at full array coverage and encounters a domain shift when applied to lower-depth nanopore data, reducing its confidence, whereas MARLIN, trained on sparse data without depth-based calibration, lacks a mechanism to attenuate confidence as sequencing depth increases. Grouping biologically distinct entities into broader methylation classes also lowers the difficulty of the classification task; because Lamprey instead separates closely related categories, its accuracy is measured against a more demanding standard.

### Lamprey reaches a confident classification within minutes of sequencing

Because nanopore methylation calls accumulate continuously during sequencing, Lamprey can reach a confident classification before run completion. Among single-plex samples (n = 174), 74.1% reached the 0.90 confidence threshold by 400 minutes and 74.7% by run completion, indicating that additional sequencing beyond this point provided minimal gain. Most confident classifications were achieved much earlier: 56.9% of samples reached threshold within 10 minutes, 62.1% within 30 minutes, and 68.4% within two hours (Fig. 5A). Because each single-plex sample receives the full output of a flow cell, these trajectories represent the fastest achievable time-to-threshold; multiplexing reduces per-sample throughput and lengthens time-to-threshold (Extended Data Fig. 6).

**Figure 5.**
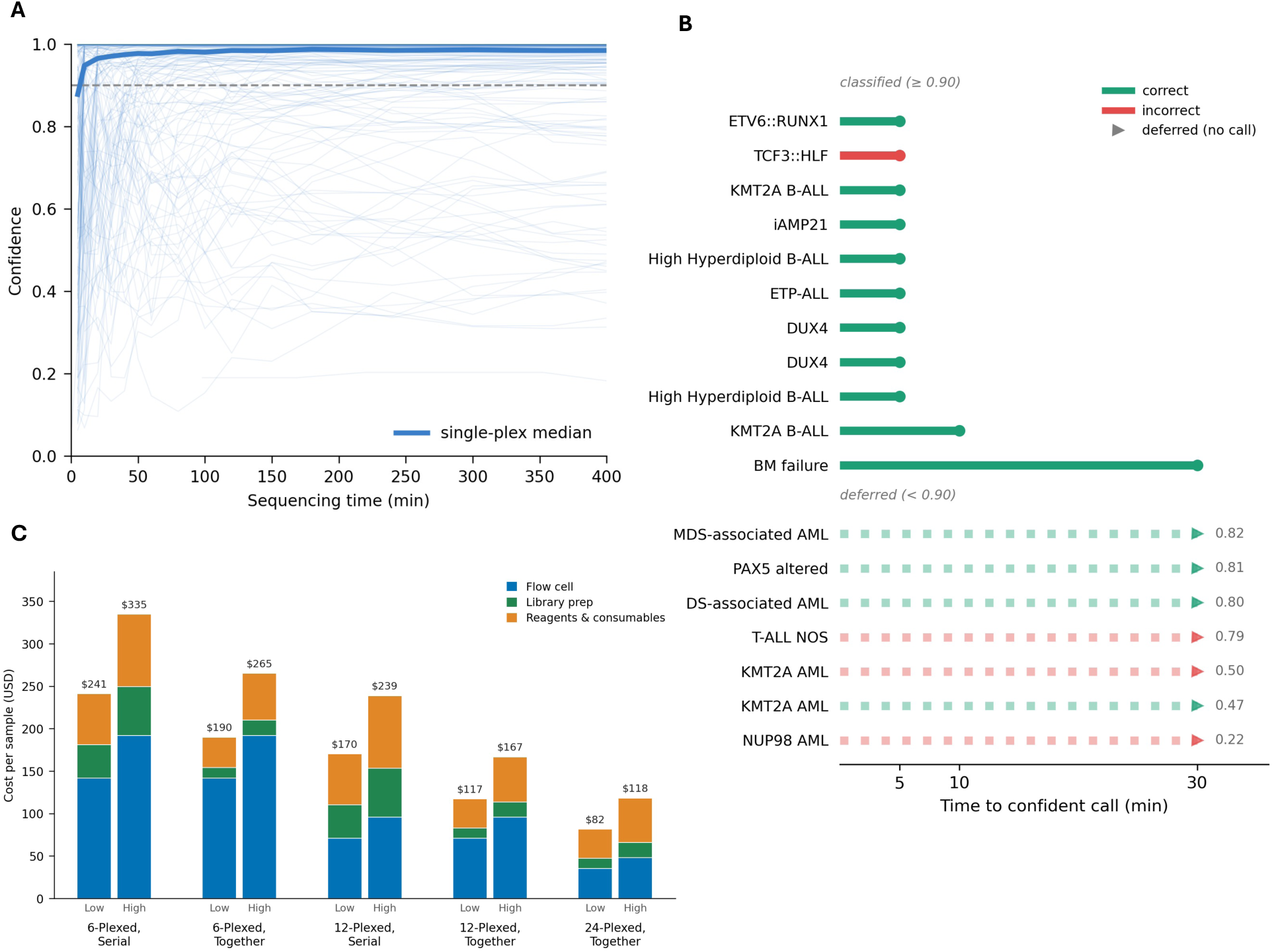
Time to diagnosis, per-sample cost, and prospective classification. **A.** Calibrated top-class confidence as a function of sequencing time for single-plex samples (n=174). Thin lines, individual sample trajectories; bold line, the across-sample median trajectory. The dashed horizontal line marks the 0.9 diagnostic threshold. **B.** Prospective classification of 18 consecutively accrued patients sequenced on the day of or the day after sample receipt, labeled by reference diagnosis. Samples reaching the 0.9 threshold (top, “classified”) are shown as lollipops whose length indicates time to confident call (minutes); green denotes agreement with the reference diagnosis and red a discordant call. Samples not reaching the threshold (bottom, “deferred”) are shown as dotted trails terminating in a triangle (no call issued), annotated with the final calibrated confidence; trail color indicates whether the top-ranked, withheld class was concordant (green) or discordant (red). **C.** Estimated per-sample cost (USD) across multiplexing levels (6-, 12-, and 24-plex) and two run configurations, serial sequencing on a shared flow cell (“Serial”) and concurrent barcoded sequencing (“Together”), with low and high estimates for each, broken down by flow cell, library preparation, and reagents and consumables. 24-plex is shown for the concurrent configuration only. Low and high estimates reflect the minimum and maximum reagent procurement prices across three participating centers (Methods). Total per-sample cost is shown above each bar.

### Per-sample cost scales with multiplexing level

We estimated reagent and consumable cost across multiplexing levels (6-, 12-, and 24-plex) and two run configurations, serial sequencing on a shared flow cell and concurrent barcoded sequencing (Fig. 5C). Per-sample cost fell from $241–335 at 6-plex serial to $82–118 at 24-plex concurrent (low and high estimates), and at matched plex costs were lower for concurrent than for serial sequencing. These figures reflect reagent and consumable costs and exclude instrument capital, labor, and computational infrastructure. Multiplexing thus yields a substantial cost reduction at the expense of time-to-diagnosis, a trade-off that can be set per sample according to clinical urgency and volume of new incidence cases. Where the same run is also used to call genomic alterations, the achievable multiplexing level is set by the higher per-sample depth that variant calling requires rather than by methylation-based classification, which remains reliable at the lowest coverage tested.^18^

### Prospective same-day classification in a routine workflow

To assess performance prospectively, we sequenced 18 consecutively accrued patients at the Princess Máxima Center, processed on the day of or the day after sample receipt and analyzed independently of all retrospective cohorts. Eleven of 18 samples (61.1%; 95% CI, 38.6 to 79.7) reached the 0.90 confidence threshold, of which 10 were correctly classified (90.9%; 95% CI, 62.3 to 98.4); consistent with the trajectories above, 9 of these 11 crossed the threshold within 5 minutes of sequencing and all within 30 minutes (Fig. 5B). Three further samples were misclassified but fell below the confidence threshold and were therefore correctly withheld from a confident call. A single confident error occurred in a *TCF3*::*HLF*-rearranged B-ALL classified as *TCF3*::*PBX1*-rearranged B-ALL (confidence 0.955); these entities share the same *TCF3* fusion partner and produce closely related methylation profiles, and *TCF3*::*HLF* is markedly underrepresented in the reference cohort (5 vs. 74 training samples). The error is nonetheless clinically consequential, as *TCF3*::*HLF* defines a high-risk leukemia whereas *TCF3*::*PBX1* carries a more favorable prognosis, and it highlights the value of the orthogonal structural-variant readout available from the same run for resolving the underlying fusion. Although limited in size, this prospective series demonstrates the feasibility of same-day molecular classification within a routine diagnostic workflow.

## Discussion

DNA methylation provided a robust molecular fingerprint for classifying hematologic malignancies across their full biological spectrum, in a single integrated readout across 86 entities. Across retrospective, external, and prospective cohorts comprising 640 patients from multiple countries, accuracy exceeded 98% among classified cases, supporting the potential for broad clinical deployment.

Current diagnostic workflows are fragmented and sequential, and the correct diagnosis is often reached only once the appropriate downstream test has been selected, creating delay even in well-resourced systems, particularly when the presentation is atypical or diagnostically overlapping. Lamprey provides a rapid molecular classification that requires no prior assumption about lineage, disease category, or suspected genetic subtype, often within minutes of sequencing and well before a full genomic readout has accumulated. This compression of the diagnostic timeline could reduce uncertainty for clinicians, patients, and families, prevent delayed treatment adaptation, and support earlier identification of patients eligible for targeted therapies or molecularly stratified clinical trials.

The implications may be greatest in low- and middle-income settings, where access to the full cytogenetic, molecular, and expert-pathology stack is limited or centralized and a substantial proportion of patients never receive a complete molecular diagnosis.^30^ A single, portable, cost-effective nanopore assay delivering rapid methylation classification alongside genomic characterization could expand access to comprehensive diagnostics rather than merely accelerate existing workflows.

The same sequencing run also yields copy-number profiles, structural variants, fusion genes, and single-nucleotide variants as depth accumulates. NASVar demonstrates that nanopore variant calling matches the performance of established short-read workflows, and together Lamprey and NASVar deliver an integrated epigenetic and genomic diagnostic readout from a single assay. This dual readout changes how a workup can be sequenced: rather than committing every patient to the full diagnostic cascade up front, Lamprey returns a confident same-day classification for the majority within the first hour, while cases that remain below the confidence are deferred to the genomic data accumulating on the same run. In well-equipped centers this combination is realistically positioned to replace much of the conventional diagnostic stack (karyotyping, FISH, PCR, RNA- and targeted sequencing, SNP arrays, and whole-genome sequencing) within a single rapid workflow, extending recent evidence that broad molecular diagnostic strategies can outperform conventional workflows while reducing costs.^31^ Because a confident classification is often reached early, sequencing can be stopped once a diagnosis is established, reducing cost, or continued to full depth for complete genomic characterization; this decision can be made per patient, balancing diagnostic sufficiency against the value of additional genomic information.

Our study has limitations. Classification performance depends on sample purity, and low-tumor-content entities frequently fell below the confidence threshold. Not all entities have been validated on nanopore data, and some are not yet represented owing to an absence of array training data. Additionally, performance for very rare entities and biologically adjacent molecular subtypes will depend on continued expansion of the reference cohort. Updated versions of Lamprey are anticipated with extended reference datasets, with the label-propagation approach allowing incorporation of new data with minimal additional curation. The prospective cohort was small, and one sample was confidently misclassified into a related entity with a different risk profile, underscoring the value of the concurrent structural-variant readout for resolving such cases. As a next step, clinical implementation studies across a variety of contexts to prove sustainability, scalability and clinical utility are required.

In conclusion, Lamprey provides a scalable, platform-agnostic framework for the molecular classification of hematologic malignancies that, combined with variant calling on the same sequencing run, could replace much of the conventional diagnostic workflow in well-equipped centers and extend comprehensive molecular diagnostics to settings where it is currently inaccessible.

**Table 1.** Characteristics of the evaluation cohorts.

| Characteristic | Retrospective | External | Prospective |
| --- | --- | --- | --- |
| No. of samples | 391 | 244 | 18 |
| No. of patients | 379 | 244 | 18 |
| Pediatric | 297 | 244 | 18 |
| Adult | 94 | 0 | 0 |
| Diagnostic category, no. |  |  |  |
| B-lineage ALL/LBL | 115 | 207 | 10 |
| AML (incl. APL) | 97 | 20 | 5 |
| T-lineage ALL/LBL | 46 | 9 | 2 |
| Mature B-cell lymphoma | 94 | 0 | 0 |
| Mature T-cell lymphoma | 12 | 0 | 0 |
| Histiocytic (LCH) | 7 | 0 | 0 |
| Mixed-phenotype acute leukemia | 1 | 1 | 0 |
| Other myeloid (CML/CMML) | 3 | 1 | 0 |
| Plasma-cell neoplasm | 1 | 0 | 0 |
| Non-neoplastic control | 15 | 6 | 1 |
| No. of distinct entities | 48 | 28 | 12 |

## Extended Data

**Extended Data Table 1.** Effect of data curation on classifier performance and high-confidence call rate.

| Configuration | Accuracy | Weighted F1 | High-conf. accuracy | % High-conf. |
| --- | --- | --- | --- | --- |
| Base dataset | 87.5% | 0.733 | 100% | 51.8% |
| PCA/model removal only | 85.7% | 0.704 | 100% | 67.9% |
| Label propagation only | 87.5% | 0.712 | 100% | 58.9% |
| PCA + label propagation (final) | 87.5% | 0.704 | 100% | 71.4% |

**Extended Data Table 2.** CpG island methylator phenotype (CIMP) status across T-ALL methylation subtypes.

| Subtype | CIMP low | CIMP high |
| --- | --- | --- |
| BCL11B | 1 | 0 |
| ETP | 0 | 20 |
| TALLOTHER | 1 | 65 |
| TALR | 65 | 5 |
| ZNF384 | 1 | 0 |
| UNCLEAR | 10 | 27 |

**Extended Data Table 3.** Depth-aware simulation and residual architecture are jointly required for optimal nanopore performance.

| Configuration | Accuracy | Weighted F1 | High-conf. accuracy | % High-conf. |
| --- | --- | --- | --- | --- |
| V1 (Sturgeon) + sparse training | 80.4% | 0.645 | 93.5% | 55.4% |
| V1 + full-range, depth-aware | 76.8% | 0.633 | 100% | 33.9% |
| V2 (ResNet) + sparse training | 83.9% | 0.690 | 100% | 69.6% |
| V2 + full-range, Bernoulli | 80.4% | 0.659 | 91.9% | 66.1% |
| V2 + full-range, depth-aware | 87.5% | 0.704 | 100% | 71.4% |

V1, original Sturgeon feedforward architecture; V2, residual network with LayerNorm and skip connections.

**Supplementary Data Table 1.** DNA methylation array training metadata and model cross-validation predictions

**Supplementary Data Table 2.** Lamprey, MARLIN, ALMA v2 per-sample retrospective results and standard-of-care diagnoses per patient

**Supplementary Data Table 3.** Serial dilution experiments and Lamprey predictions

**Supplementary Data Table 4.** Prospective cohort per-sample results

**Extended Data Fig. 1:**
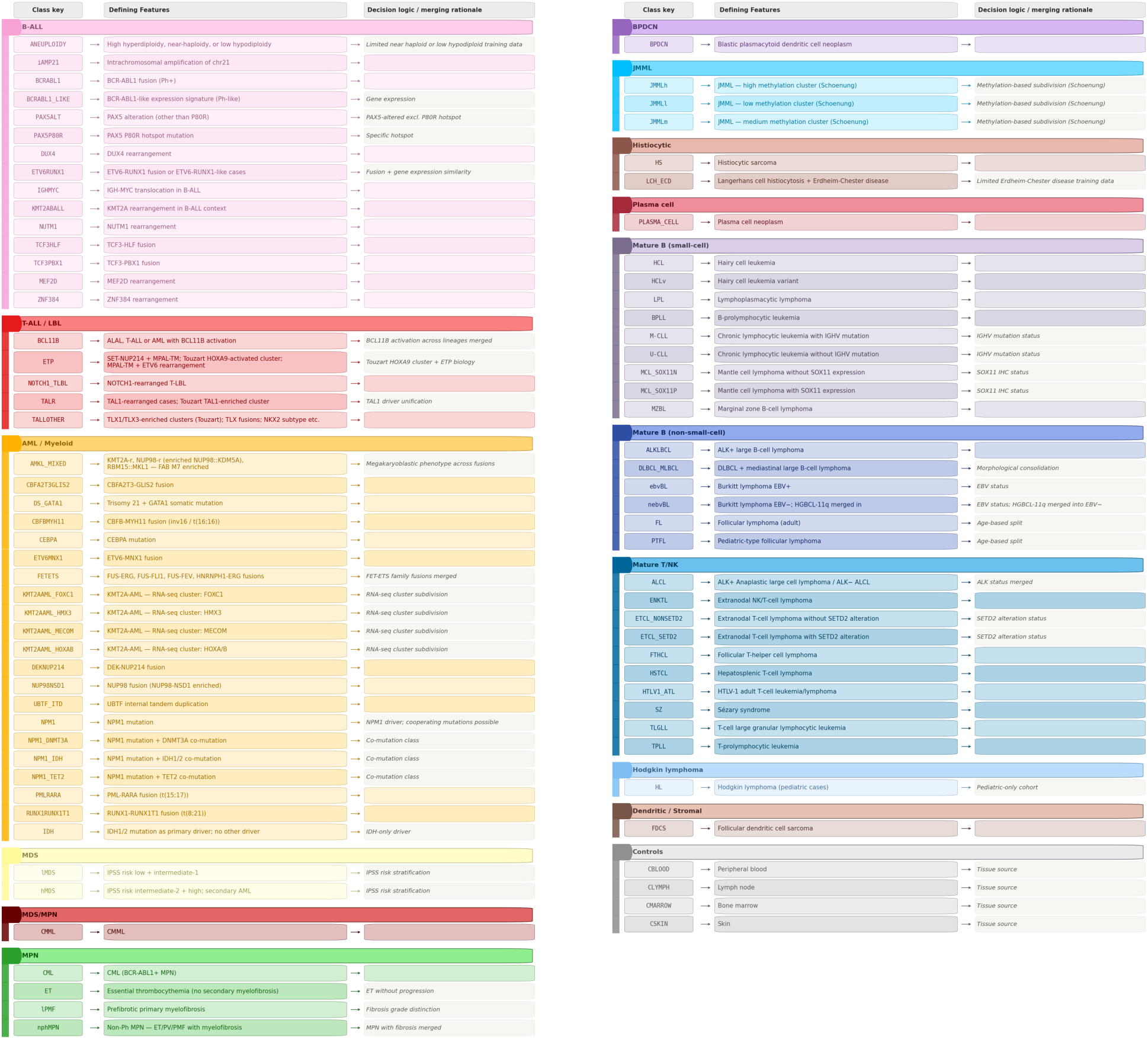
Decision logic for methylation class definitions. Table showing the name of methylation classes (left column), the defining features of the methylation class (center column) and the corresponding decision logic (right column).

**Extended Data Fig. 2:**
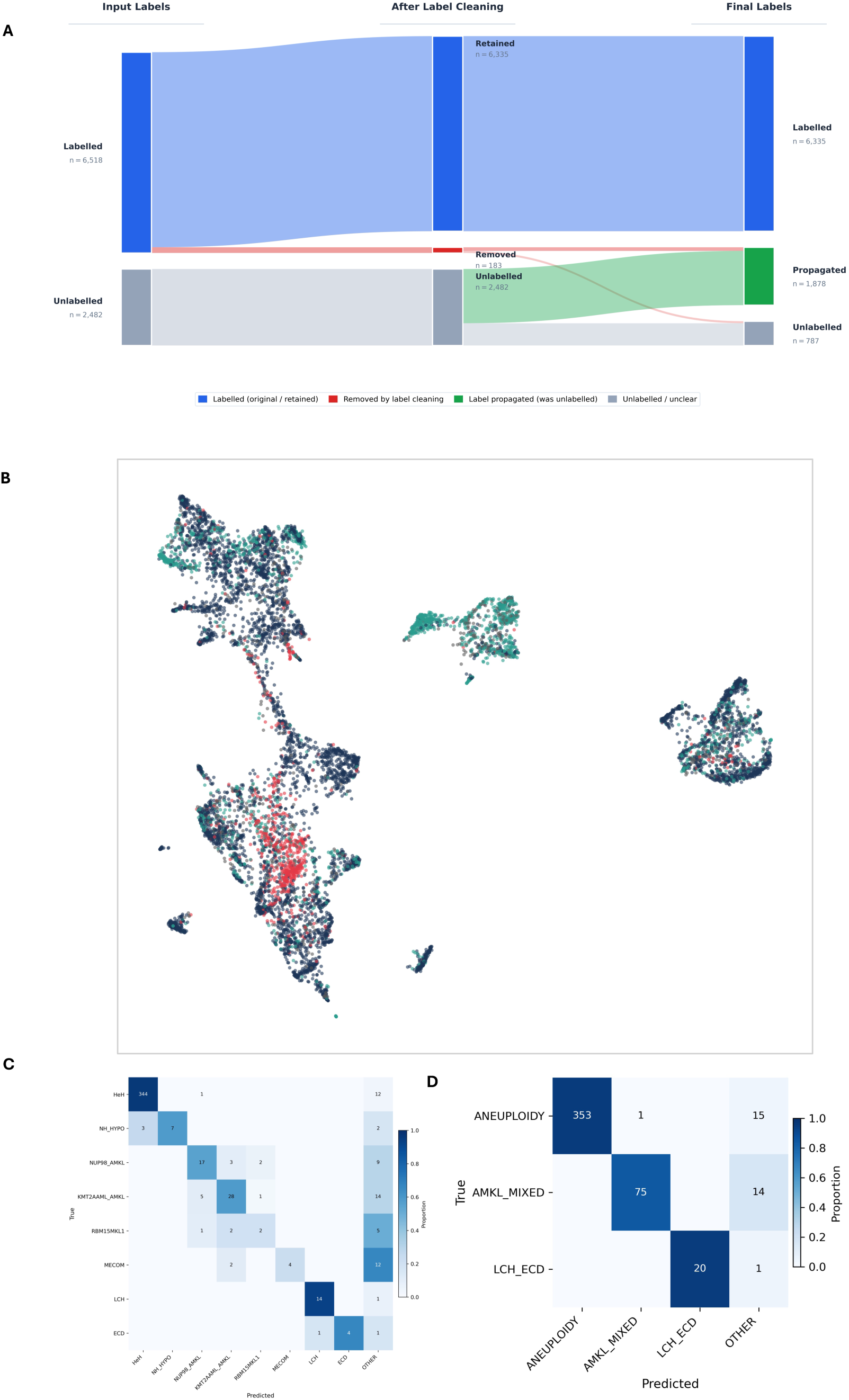
Reference-cohort curation and class merging. A) Sankey diagram illustrating the three-stage label cleaning and propagation pipeline. In stage 1, samples were assigned initial labels based on genomic and phenotypic data from their respective source publications. In stage 2, a combined PCA-based neighborhood concordance and cross-validation confidence filtering step was applied to remove samples with inconsistent or uncertain labels. In stage 3, previously unlabelled samples were assigned pseudo-labels based on high-confidence cross-validation predictions from the cleaned dataset. B) UMAP dimensionality reduction, with samples colored on whether the label assigned in the original dataset was kept (dark blue), whether the sample had no label and was labelled by propagation (green), whether the original label was removed (red) or whether the sample never received a label (grey). C) Confusion matrix showing Lamprey predictions for methylation subtypes that underwent curation during dataset preparation, evaluated on the base dataset prior to label cleaning or propagation. Subtypes shown include those that were removed from the final taxonomy (MECOM) and those that were merged: high hyperdiploidy with near-haploid and low-hypodiploid BCP-ALL (HHYPERDIPLOID with NH_HYPO), NUP98-rearranged and KMT2A-rearranged AMKL (NUP98_AMKL and KMT2A_AMKL), RBM15::MKL1-rearranged AML, and Langerhans cell histiocytosis with Erdheim-Chester disease (LCH with ECD). Samples predicted as none of these subtypes are assigned to OTHER. D) Confusion matrix of methylation subtypes from the base dataset after merging of methylation classes, confirming that the merged classes are captured as unified entities by the classifier.

**Extended Data Fig. 3:**
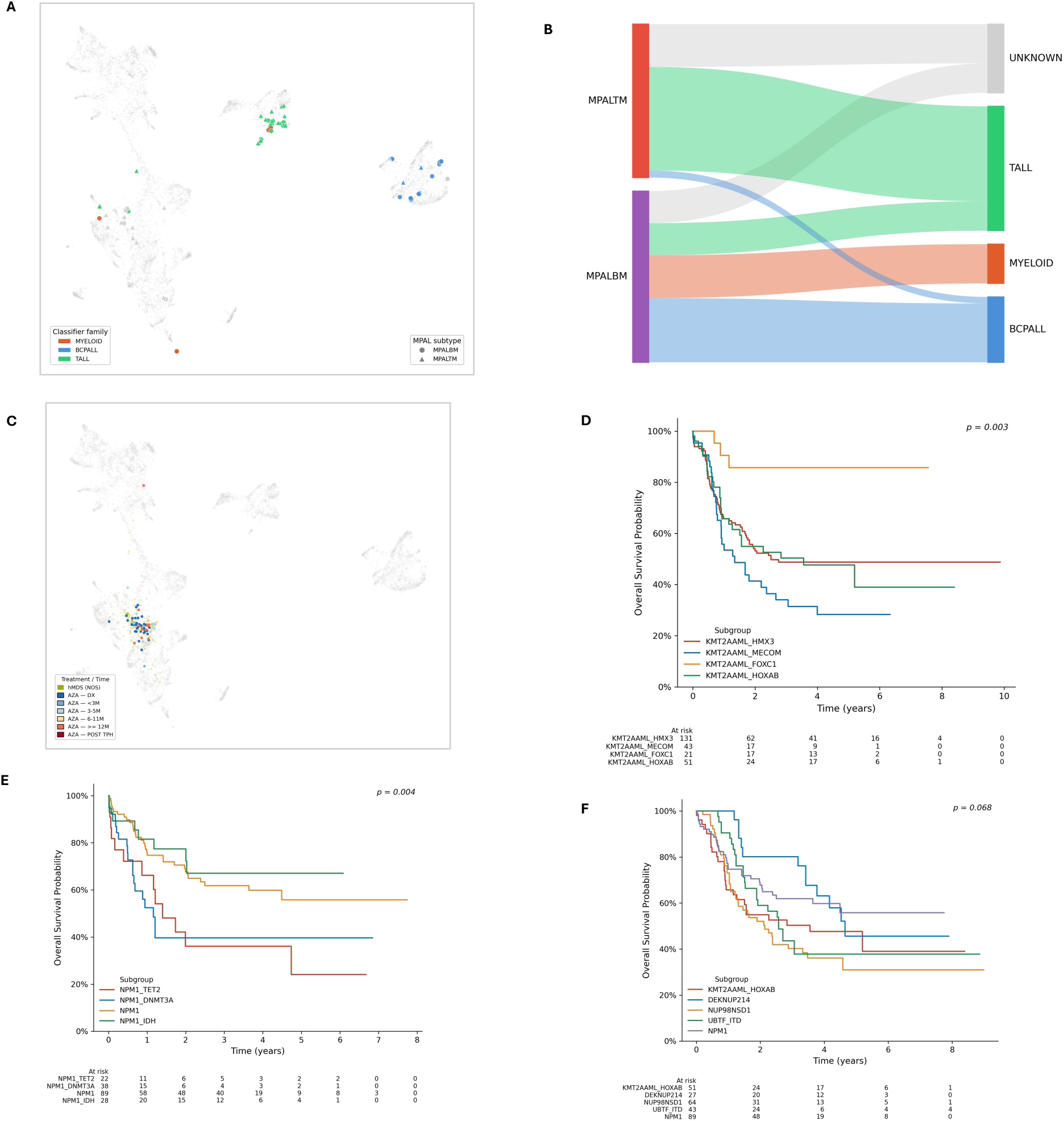
Methylation classification reflects underlying disease biology. A) UMAP dimensionality reduction of the full Lamprey training dataset with MPAL samples highlighted, colored by methylation family assignment. Methylation family scores were derived by aggregating Lamprey subtype scores within each family, with a minimum score of 0.9 required for a family-level classification to be assigned. Samples not reaching this threshold are shown as unclassified. B) Sankey diagram showing the distribution of methylation family assignments stratified by immunophenotypic MPAL subtype (T/Myeloid, B/Myeloid). Samples for which no methylation family reached a score of 0.9 are classified as unknown. C) UMAP dimensionality reduction of the full Lamprey training dataset with high-risk myelodisplastic samples highlighted, colored by azacitidine (AZA) treatment timepoint. Samples with no treatment information are labeled hMDS (NOS). D) Kaplan-Meier overall survival curves for KMT2A-rearranged AML subtypes E) Kaplan-Meier overall survival curves for NPM1-rearranged AML subtypes F) Kaplan-Meier overall survival curves for HOXA/B activated AML subtypes. Subtype labels reflect annotations derived from the original dataset. Log-rank p-values shown in each panel.

**Extended Data Fig. 4:**
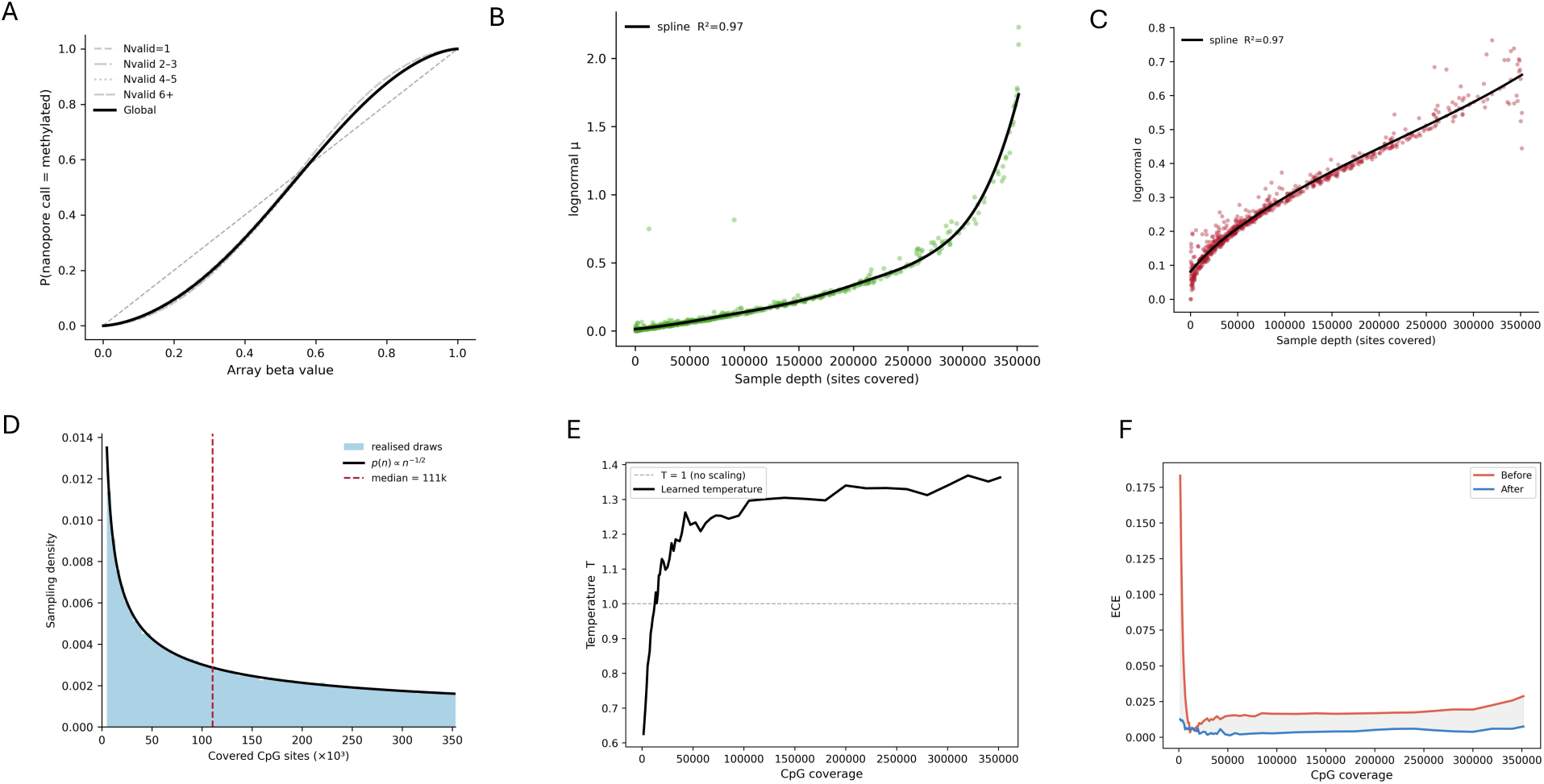
Empirical models underlying the array-to-nanopore simulation and depth-aware calibration. A) Logistic calibration curves relating array beta values to the probability of a nanopore methylation call of 1, fitted globally (black) and within read per CpG site (Nvalid) strata (grey). The S-shaped deviation from the identity line (dashed) reflects systematic caller bias independent of read depth. B) Per-sample lognormal μ parameter of the Nvalid distribution, governing the mean of the log-transformed per-site read depth, where higher values correspond to more reads per CpG site on average. Plotted against sample depth (sites covered), with cubic smoothing spline fit (black, R² = 0.97). Each point represents one nanopore sample (n = 733).Per-sample lognormal σ parameter of the Nvalid distribution plotted against sample depth, with cubic smoothing spline fit (black, R² = 0.97). C) Per-sample lognormal σ parameter of the Nvalid distribution, governing the spread of the log-transformed per-site read depth, where higher values indicate greater variability in read depth across CpG sites within a sample. Plotted against sample depth, with cubic smoothing spline fit (black, R² = 0.97). Each point represents one nanopore sample (n = 733). D) Inverse-square-root sampling density of the per-iteration CpG count. For each training iteration, the number of covered CpG sites was drawn from [5,000; 353,232] with probability proportional to n(−1/2), upweighting low-coverage samples so the model trains across the full sparsity range (median ≈ 111k). The histogram shows 300,000 realized draws; the black curve is the analytic density. E) Post-hoc temperature scaling parameters learned per adaptive CpG coverage bin. Values above 1 indicate overconfidence (softening required); values below 1 indicate underconfidence. The dashed line marks T = 1 (no correction). F) ECE computed within each CpG coverage bin before (red) and after (blue) depth-aware temperature scaling. The shaded region highlights the reduction in miscalibration across the coverage spectrum.

**Extended Data Fig. 5:**
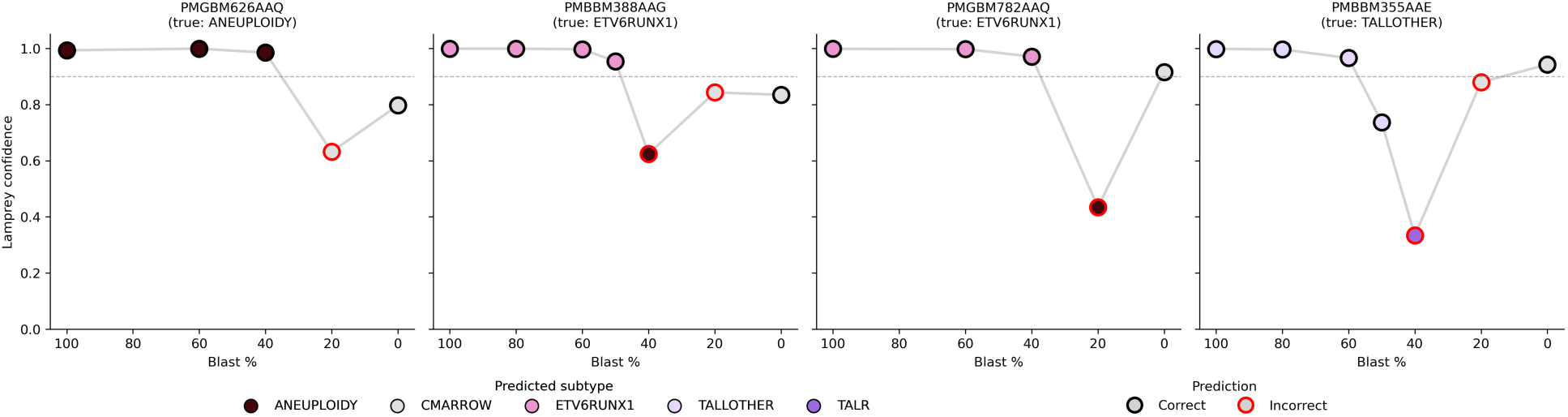
Effect of tumor purity on classification confidence. Lamprey classification across a blast-cell dilution series. Bone marrow aspirates from leukemia cases were serially diluted with matched normal bone marrow (CMARROW) to generate a range of tumor purities, and each dilution was profiled by nanopore sequencing and classified with Lamprey. Each panel shows one dilution series, with blast percentage on the x-axis (decreasing left to right) and Lamprey’s predicted-class confidence on the y-axis. Points are colored by the predicted subtype (legend), and edge color indicates whether the prediction matched the expected call (black = correct, red = incorrect); at 0% blasts, the expected call is CMARROW. The dashed horizontal line marks the 0.9 high-confidence threshold. Panel titles indicate the sample identifier and its true molecular subtype.

**Extended Data Fig. 6:**
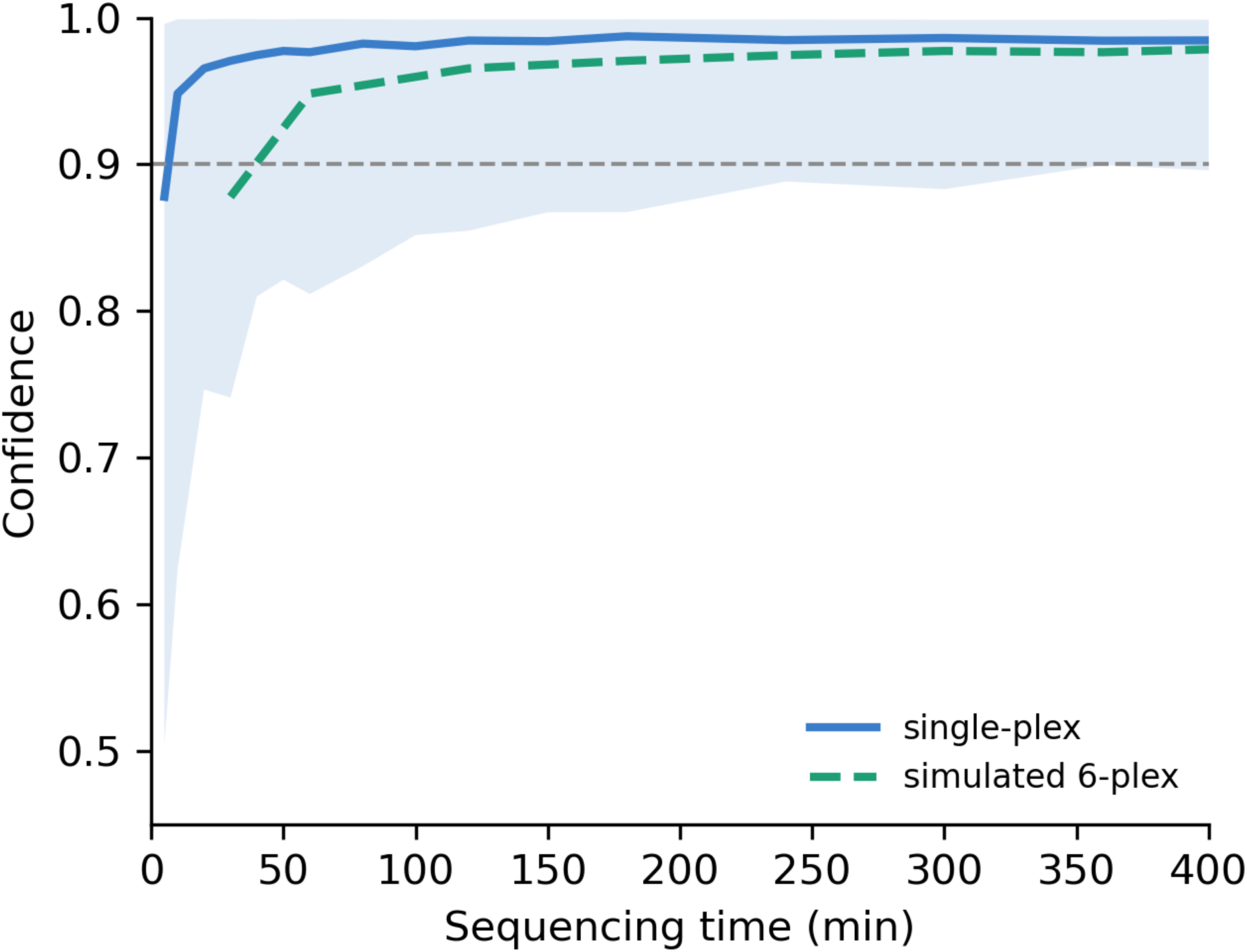
Time to confident classification across multiplexing configurations. Calibrated top-class confidence as a function of sequencing time for single-plex runs. The solid line shows the median single-plex trajectory and the shaded band the interquartile range (25th–75th percentile). The dashed line shows a simulated six-plex configuration, obtained by rescaling the single-plex time axis by a factor of six to approximate the ∼6-fold reduction in per-sample throughput when six barcoded samples share a single flow cell; it therefore reflects the same per-sample read accumulation displaced in wall-clock time rather than an independently sequenced multiplexed run. The horizontal dashed line marks the 0.9 diagnostic threshold.

